# Risks to Children under-five in India from COVID-19

**DOI:** 10.1101/2020.05.18.20105239

**Authors:** Isabel Frost, Katie Tseng, Stephanie Hauck, Geetanjali Kapoor, Aditi Sriram, Arindam Nandi, Ramanan Laxminarayan

## Abstract

**Objective:** The novel coronavirus, COVID-19, has rapidly emerged to become a global pandemic and is known to cause a high risk to patients over the age of 70 and those with co-morbidities, such as hypertension and diabetes. Though children are at comparatively lower risk compared to adults, the Indian population has a large young demographic that is likely to be at higher risk due to exposure to pollution, malnutrition and poor access to medical care. We aimed to quantify the potential impact of COVID-19 on India’s child population.

**Methods:** We combined district family household survey data with data from the COVID-19 outbreak in China to analyze the potential impact of COVID-19 on children under the age of 5, under three different scenarios; each of which assumed the prevalence of infection to be 0.5%, 1%, or 5%.

**Results:** We find that in the lowest prevalence scenario, across the most populous 18 Indian states, asymptomatic, non-hospitalized symptomatic and hospitalized symptomatic cases could reach 87,200, 412,900 and 31,900, respectively. In a moderate prevalence scenario, these figures reach 174,500, 825,800, and 63,800, and in the worst case, high prevalence scenario these cases could climb as high as 872,200, 4,128,900 and 319,700.

**Conclusion:** These estimates show COVID-19 has the potential to pose a substantial threat to India’s large population of children, particularly those suffering from malnutrition and exposure to indoor air pollution, who may have limited access to health services.

## Main Text

## Introduction

The novel coronavirus, COVID-19, has spread from Wuhan, China to become a worldwide pandemic. Studies from Wuhan indicate that the highest risk categories are adults over the age of 80 and those with comorbidities, such as uncontrolled hypertension, diabetes, cardiovascular disease, chronic lung disease, and cancer [1]. Although the documented short-term risk posed by COVID-19 to children is lower than in adults, it is possible that they may present a sizeable burden in a country such as India that is home to 130 million children under the age of five, many of whom are undernourished, suffer from non-COVID19 respiratory illnesses and have risk factors for respiratory illness, including household air pollution and exposure to second-hand tobacco smoke [2-4].

Already, every year in India 169,760 deaths of children under 5 from pneumonia could be averted through prompt access to an appropriate antibiotic [5]. This situation, of low access to essential healthcare, stands to be further exacerbated by higher demands on the health system due to COVID-19. Seasonal peaks in acute respiratory infections are likely to place further demands on the Indian health system, for example in Pune respiratory syncytial virus cases peak during the rainy season in July [6]. Children living in India are more vulnerable to severe respiratory infections due to high rates of pollution, indoor air pollution [2-4], undernutrition [7-9], crowding, lack of exclusive breastfeeding, a low degree of maternal education, limited access to secondary care and passive health seeking behaviors [10-11]. Socio-economic status determines housing size and lower income households are more crowded, increasing the risk of pathogen exposure and transmission. Crowding is also linked to women’s education as educated women tend to have fewer children, leading to less crowding. Cooking indoors with open fires, unprocessed biomass, and inadequate ventilation generates high levels of indoor air pollution. Chemicals in wood smoke provoke inflammatory responses in the lungs, irritate airways and suppress the immune system via the macrophage response.

Mounting evidence from hotspots of the pandemics suggests fewer children show severe symptoms of COVID-19. Only 1% of 72,314 cases in China were less than ten years of age [12] and in an analysis of 2,143 Chinese pediatric patients, only one child died [13] while in Italy none of 1,652 deaths were under 30 years of age [14]. 5.9% of child cases were severe or critical, in contrast to 18.5% of adult cases [13]. The risk of symptomatic infection has also been reported to be lower in pediatric populations [15]. In the largest study of pediatric COVID-19 patients to date, 94.1% of 2,143 patients under the age of 18 reporting to the Chinese authorities were asymptomatic, mild or moderate cases[13]. However, of the younger age group, infants were particularly vulnerable to infection, with 10.6% of severe and critical cases under the age of one.

Children are less likely to suffer from the comorbidities associated with high susceptibility to COVID-19 in adult populations, including Type 2 diabetes, heart disease, hypertension, autoimmune diseases, and chronic kidney disease. Unfortunately, the data currently available from child cases do not include the majority of risk factors, such as chronic respiratory conditions, use of steroids, hemodynamically significant heart disease (CHD), chronic kidney disease, or other immunosuppressive syndromes, such as TB or HIV. Therefore, based on current evidence it is unclear how children with comorbidities will respond to more severe forms of COVID-19. A case report from 2017 on HCoV-NL63 in a malnourished infant demonstrated that when comorbidities are present, normally benign coronavirus infections can lead to severe disease and death within 24 hours.

Irrespective of the presence of comorbidities, there may also be mechanistic reasons for differences in susceptibility to COVID-19 between adults and children. Evidence suggests the Angiotensin converting enzyme II (ACE2) may be the cell receptor used by COVID-19 to bind and some have suggested the maturity and function of this protein may be less developed in children [13].

## Methods

Current evidence suggests that children are at lower risk from severe disease. However, given India’s large young population, the need for hospital beds and critical care is likely to be high. Here we present the data for infections in children (aged 0-4) under three different scenarios given COVID-19 prevalence rates of 0.5% (Figure 1), 1% (Figure 2), or 5% (Figure 3). The figures depict only the largest states in India due to a wide range in state population sizes. The age structure of the Indian population was taken from the most recent National Family Health Survey data (NFHS-4, 2015-2016) for India [16].

**Figure 1.**
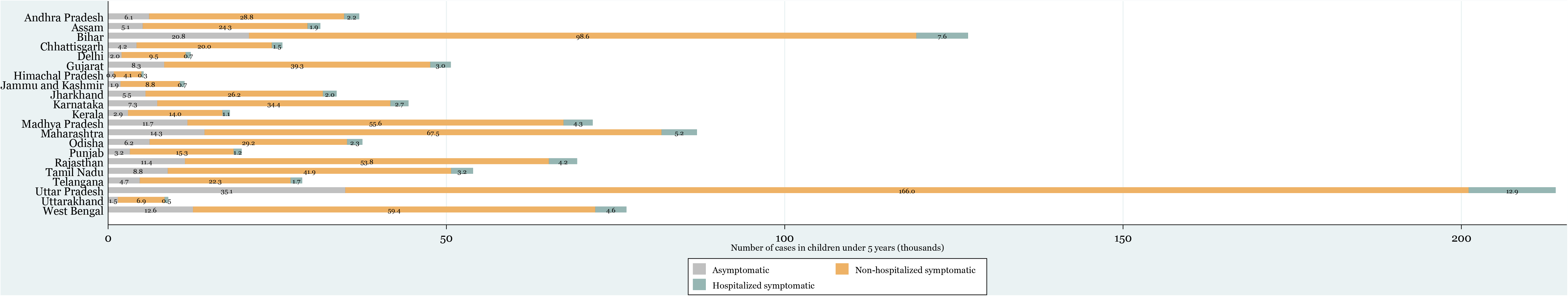
Number of cases in children under 5 years old given infection rate of 0.5%.

**Figure 2.**
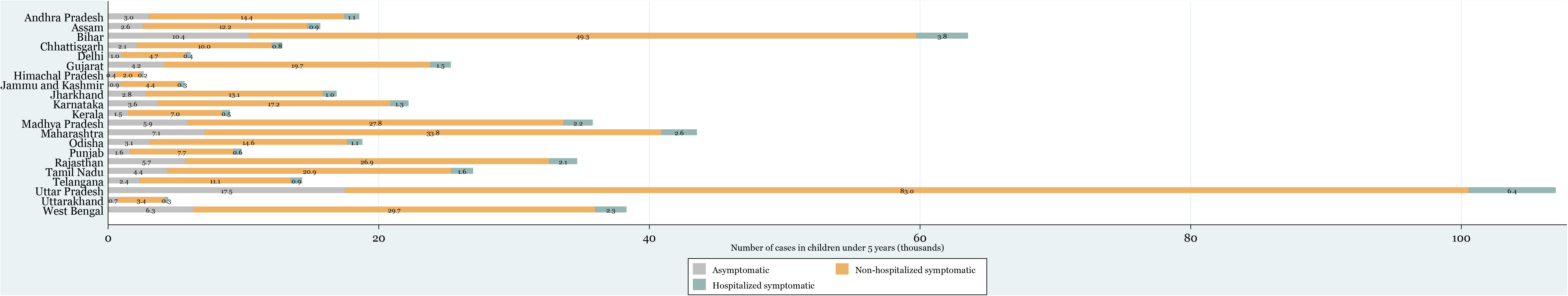
Number of cases in children under 5 years old given infection rate of 1%.

**Figure 3.**
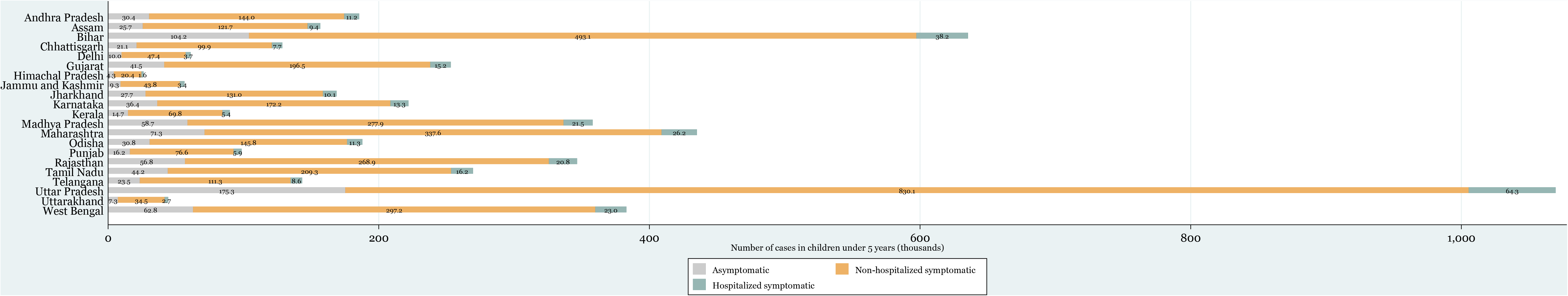
Number of cases in children under 5 years old given infection rate of 5%.

The asymptomatic and symptomatic rates used in the calculations were inferred from Liu *et al*. [17] based on the mean rates and standard deviations reported for each level of disease severity for those under 65 years of age. Of those infected with COVID-19, 16.39% were assumed to be asymptomatic or to have mild symptoms and symptomatic rates in this age group were 83.61%. The hospitalization rate of 6.01% was also drawn from Liu *et al*. [17] and assumed that severe and critically ill symptomatic cases are hospitalized, while moderate symptomatic cases are not hospitalized. The following definitions of levels of disease severity were used [17]; asymptomatic patients showed no clinical symptoms but returned a positive nucleic acid test; mild illness involved mild clinical symptoms but without evidence of pneumonia; moderately ill patients had a fever, showed respiratory symptoms, and imaging showed pneumonia; severely ill patients had one of the following: respiratory distress, defined as a respiratory rate greater than or equal to 30 beats per minute; a mean oxygen saturation in resting state that was less than or equal to 93%; or arterial blood oxygen partial pressure less than or equal to 300 mmHg (1 mmHg = 0.133kPa); critically ill patients displayed one of the following: respiratory failure requiring mechanical ventilation; shock; combined organ failure requiring ICU monitoring and treatment.

## Results

Considering a low COVID-19 prevalence of 0.5%, the 18 largest Indian states would be likely to collectively suffer 31,900 children hospitalized with COVID-19 (Figure 1). The effect is likely to be greatest in the most populous states, such as Uttar Pradesh which could have 6,400 hospitalized symptomatic cases, and 83,000 non-hospitalized symptomatic cases in children under the age of 5. Also worrying, in the state of Uttar Pradesh alone there would be 17,500 asymptomatic cases of COVID-19 in children, with the potential to spread this to more vulnerable age groups. At higher COVID-19 prevalence rates of 1% (Figure 2) and 5% (Figure 3), the 18 largest states could reach 63,800 and 319,700 in the number of hospitalized child cases respectively. Under the scenario with the highest prevalence rate we explored, of 5%, Uttar Pradesh could have 64,300 hospitalized, 830,100 symptomatic and 175,300 asymptomatic cases of COVID-19 in children under 5 years of age alone.

## Discussion

Of 72,314 cases documented by the Chinese Centers for Disease Control, only 1% of patients were under the age of 10. Of those, 12.3% of children tested positive with a median age of infection of 6.7 years [12]. Of 2,143 Chinese pediatric COVID-19 patients, only one child died and most cases were mild, with much fewer severe and critical cases (5.9%) than adult patients (18.5%) [13]. Of 22,512 people in Italy with COVID-19, there were 1,625 deaths but none of these were in people younger than 30 years [14]. The fact that 15% of children overall were asymptomatic in China and had no radiological findings suggests many children could be asymptomatic carriers of the novel coronavirus [12]. According to our calculations, even in the scenario with the lowest prevalence rate of 0.1%, the total number of asymptomatic carriers of COVID-19 under the age of 5 was 87,200, and in the high prevalence scenario of 5% this reached 872,200 in the 18 most populous Indian states alone.

In the largest study of pediatric COVID-19 patients to date, of 2,143 patients under the age of 18 reporting to the Chinese authorities between January 16 and February 8, 2020, 94 (4.4%), 1,091 (50.9%) and 831 (38.8%) patients were diagnosed as asymptomatic, mild or moderate cases, accounting for 94.1% of the total cases [13]. Approximately 50% of children infected with COVID-19 exhibit high fever and/or cough with fever subsiding within three days [18]. Myalgia, lethargy, and gastrointestinal symptoms have also been reported. Only one death was reported in this period of a 14 year-old boy. Infants were particularly vulnerable to infection with 10.6% of severe and critical cases being under the age of one [13]. Children of all ages were susceptible with no significant difference between genders. 5.9% of child cases were severe or critical, in contrast to 18.5% of adult cases [13]. However, in the Indian context, given its large young population, 31,900 hospitalized cases in the low prevalence scenario, or 319,700 hospitalized cases in the high prevalence scenario would present a severe burden to the health system if these cases were to arise around the same point in time. An inundated healthcare service would be less able to provide quality care and this could cause mortality rates to increase.

Much of the data on the impact of COVID-19 on pediatric populations comes from the original epicenter of the outbreak in Wuhan, China. However, many schools were closed for the Spring Festival Holiday at this time, potentially reducing the transmission of the virus in children [19]. In early January, among 366 children (≤16 years of age) hospitalized with respiratory illness at Tongji Hospital in Wuhan, 23 had influenza A virus, 20 had influenza B virus and 6 patients were infected with COVID-19 [20]. All 6 recovered with only one being admitted to ICU and receiving pooled immune globulin. COVID-19 patients were treated empirically with antiviral agents, antibiotic agents, and supportive therapies and their recovery time was between 5 and 13 days. However, it is hard to predict the recovery trajectory these patients would have seen without access to this high level of care. Many in India cannot afford access to medical year and out-of-pocket health expenditures are estimated to push 57 million Indians into poverty each year [21].

Of the 661 COVID-19 patients at one hospital in China between Jan 17 and March 1, 36 (5%) were children (1-16 years of age) [19]. Pediatric patients acquired COVID-19 either through close contact with infected family members (89%), exposure to endemic areas (33%), or both (22%). In India, families often cohabit between multiple generations and though government mandated social distancing attempts to reduce transmission many indians have no option but to share a small co-living space. The majority (53%) of pediatric patients showed moderate symptoms and none of the cases were severe or critical. Though symptoms of COVID-19 are often mild in children, the prevalence of pneumonia with COVID-19 (53%) in this study was higher than H1N1 influenza (11%), and similar to SARS (65%) [19].

In conclusion, COVID-19 poses a rapidly emerging threat, not only to the most publicized at-risk groups; the elderly and immunocompromised, but also to younger age groups. India has a large population of children, many of whom do not have access to essential health services or are vulnerable to respiratory pathogens due to pollution-induced lung damage and malnutrition. In addition, the simple estimates presented here are unable to take into account the further impact of the reduction in services offered by an overburdened health system on children suffering from health issues other than COVID-19. Many children stand to be affected by the coming pandemic and it is essential that India prepares for this eventuality or the lives of the next generation will be put at risk.

## Key Messages

### What this study adds?

This study shows the potential of COVID-19 to pose a substantial threat to India’s large population of children, particularly those made all the more vulnerable by malnutrition and exposure to indoor air pollution, for whom access to health services is likely to be limited.

## Data Availability

Data used in the study is available from public databases as listed in the methods section of the paper.

## Authors contributions

IF, SH, AN and RL all contributed to the writing of the manuscript. KT, GK and AS performed the data collation and analysis. KT designed and prepared the figures.

